# COVID-19 infection and hospitalization according to the burden of chronic noncommunicable diseases in Brazil

**DOI:** 10.1101/2021.05.03.21256532

**Authors:** Fabiana R. Ferraz, Wolney L. Conde, Isabela Venancio, Larrisa Lopes, Catarina M. Azeredo

## Abstract

Chronic diseases, worse socioeconomic conditions and old age can increase infection and hospitalization rate due to Coronavirus disease (COVID-19). We assessed the association between the burden of NCDs and the occurrence of infections and hospitalizations of COVID-19 in Brazil in a large COVID-19 national survey data. We analyzed only data collected between July and November 2020 (*n* = 1,071,782). The frequencies of positive COVID-19 diagnosis and NCD burden were estimated according to age, sex, socioeconomic strata and skin color categories. We estimated hazard ratios and 95% confidence intervals using Cox regression models. There is a non-linear dose-response inverse association between *per capita* income and the rates of infection and hospitalization due to COVID-19. The presence of NCDs was associated with a higher incidence of COVID-19 infection (HR_1NCD_ = 1.34; 95% CI: 1.26; 1.43; HR_2 or more NCD_= 1.54 95% CI: 1.39; 1.71) and incidence of hospitalization (HR_1NCD_ = 3.08 95% CI: 2.26; 4.19; HR _2or more NCD_= 6.81 95% CI: 4.88; 9.49).The difference between the risks of infection or hospitalization of COVID-19 attributable to the burden of NCDs is non-linearly associated with the income.

## Text

Worldwide, over 93 million cases and over 2 million deaths have been reported since the start of the Coronavirus disease (COVID-19) pandemic by the World Health Organization (WHO). Brazil has reported the highest number of new cases, as of 16 March 2021 (1).

According to the WHO, 15% of COVID-19 cases are likely to develop into hospitalizations and 5% require treatment in intensive care units (ICU) (2). Factors such as comorbidities, the presence of chronic diseases, worse socioeconomic conditions and old age can increase the risk of hospitalization (3). According to data from the *Pesquisa Nacional de Saúde* (National Health Survey) (PNS 2019), approximately 52% of Brazilian adults have at least one chronic non-communicable disease (NCD). Respiratory and cardiac diseases, diabetes and hypertension are the comorbidities leading towards the worst prognosis for COVID-19 (4,5).

In Brazil, considering the prevalence of NCDs, nationwide estimates for adults at risk of severe COVID-19 range from 34.0% (53 million) to 54.5% (86 million) (6). To date we have no evidence that the presence of comorbidities increases the risk of infection or if there is a gradient between the number of comorbidities and the risk of hospitalizations due to COVID-19 (7). Therefore, the aim of this study was to analyze the association between the burden of NCDs and the occurrence of infections and hospitalizations of COVID-19.

## METHODS

### Data

We used data from PNAD COVID-19 (Portuguese acronym for National Household Sample Survey COVID-19), conducted by the Instituto Brasileiro de Geografia e Estatística (IBGE) and the Brazilian Ministry of Health. PNAD COVID-19 survey followed, through monthly telephone calls, approximately 193,000 households between May and November 2020. The sample was selected from an elaborated registry of probabilistic matching between addresses of households contained in the Brazilian National Household Sample Survey (PNAD 2019) and addresses of six databases with telephone information. The procedures, databases used, questionnaires and sample plan of the research are described in technical documents from IBGE (8). For the present study, we analyzed only data from individuals between 20 and 59 years of age, collected between July and November 2020 (n=2.650.459).

PNAD COVID-19 collected information about exams, results of COVID-19, income and sociodemographic characteristics of residents. It was also asked about previous diagnosis of diabetes mellitus, high blood pressure, chronic respiratory diseases (asthma, bronchitis, emphysema and others), heart diseases (infarction, angina, heart failure, arrhythmia), depression and cancer. Three questions related to the tested specimen were also used to diagnose COVID-19 infection, without discriminating the test performed (molecular biology, antigen or antibody). The oronasal swab test uses the RT-PCR technique to identify viral RNA, usually up to the 8th day of infection (9,10). In the venous blood test it is possible to detect antigen (current infection) or antibody (current or past infection), and it is usually applied from the 8th day after the beginning of the symptoms . The capillary blood test, also available for diagnosis, allows verification of results after 15 minutes of the exam (11). We used three diagnostic outcomes: positive diagnosis in oronasal swab, positive diagnosis in venous blood, and positive diagnosis in capillary blood.

Sociodemographic information was also analyzed, such as sex (male and female), age (in years), skin color (“white”/“yellow” and “black”/“brown”/“indigenous”), information on per capita household income (PCI), which was calculated by the monthly average of individual incomes divided by the total number of individuals in the household and then divided into fifths of the distribution; schooling, complete or not, which was grouped into the following levels: no schooling, elementary school, high school, higher education and post graduation; geographic region and area of residence (urban or rural). Predicted values adjusted for NCD, age, sex, education, income and skin color were created for two probabilistic outcomes, diagnosis and hospitalization by COVID-19.

### Statistical Analysis

For the present analysis, we carried out the following preparation: a) selection of individuals between 20 and 59 years old; b) we consider any positive result reported in any of the 3 diagnostic tests asked; c) noncommunicable Diseases (NCD) diagnoses were grouped into 0, 1, 2 or more diseases to represent the NCD burden; d) we selected all cases with complete response for variables under analysis (n = 1,071,782).

The frequencies of positive COVID-19 diagnosis and NCD burden were estimated according to age, geographic and socioeconomic strata and skin color categories. We calculated hazard ratios (HR) and 95% confidence intervals (CI) for the association between NCD and the risk of infection and hospitalization for COVID-19 using Cox regression models via maximum likelihood to account for repeated measures, as each person could contribute with one or more interviews to the study. Findings at p < 0.05 were considered statistically significant. Initially, we assessed the number of chronic noncommunicable diseases in separate models without including any covariates.. Secondly, we included a priori selected demographic variables to assess potential confounding: sex, age, race/skin color, education, household income and rural/urban residency. The adjusted prevalence of COVID-19 was predicted according to NCD burden and stratified by per capita household income. We estimated p-trends for the dose-response relationship between NCD and COVID infection and hospitalization. Finally, predicted values from the model were used to estimate COVID-19 latent frequency according to the burden of NCD as grading risk to the infection or ICU use. All analyses were performed using sampling weights in Stata® 15.1.

## RESULTS

The prevalence of COVID-19 was higher among individuals aged 30 to 50 years (58%), while the prevalence of hospitalizations was higher among individuals aged 40 or over more (59%). Around 60% of the people in the age group of 40 years or over had NCDs. The prevalence of 2 or more NCDs among the richest was 1.3 times the prevalence among the poorest, however, the prevalence of hospitalizations was 1.2 times more frequent in the poorest group.

The COVID-19 infection rate as a function of the NCD burden was not statistically associated with sex. Table 2 shows the association between NCD burden and outcomes of COVID-19. The presence of NCDs was associated with a higher incidence of COVID-19 infection for 1 NCD (HR = 1.34; 95% CI: 1.26; 1.43) and for 2 or more (HR = 1.54 95% CI: 1.39; 1.71). Also, the presence of NCDs was associated with higher incidence of hospitalization for 1 NCD (HR = 3.08 95% CI: 2.26; 4.19) and for two or more NCDs (HR = 6.81 95% CI: 4.88; 9.49).

**Table 1.** Prevalence of Chronic Noncommunicable Diseases and gross probability of positive diagnosis and hospitalization for COVID-19. Brazil PNAD COVID-19, 2020.

| Characteristics | Noncommunicable Diseases (%) |  |  | COVID-19 (%) |  |
| --- | --- | --- | --- | --- | --- |
|  | None | 1 | 2 or more | Confirmed cases | Hospitalization |
| **Brazil** | 79.2 | 16.0 | 4.8 | 3.2 | 6.8 |
| **Age** |  |  |  |  |  |
| 20 - 30 | 30.9 | 13.7 | 4.7 | 23.8 | 16.1 |
| 30 - 40 | 29.1 | 19.8 | 12.2 | 30.3 | 24.9 |
| 40 - 50 | 24.0 | 30.4 | 30.4 | 27.3 | 30.1 |
| 50 - 59 | 16.0 | 36.1 | 52.7 | 18.6 | 28.9 |
| **Gender** |  |  |  |  |  |
| Male | 49.5 | 41.3 | 34.1 | 44.2 | 46.2 |
| Female | 50.5 | 58.7 | 65.9 | 55.8 | 53.8 |
| **Geographic Region** |  |  |  |  |  |
| N | 13.1 | 9.7 | 8.1 | 20.4 | 14.3 |
| NE | 30.8 | 28.6 | 26.4 | 31.6 | 26.2 |
| SE | 28.6 | 32.0 | 33.1 | 22.1 | 24.9 |
| S | 16.8 | 18.5 | 20.7 | 11.3 | 16.5 |
| CW | 10.7 | 11.2 | 11.7 | 14.6 | 18.1 |
| **Area** |  |  |  |  |  |
| Urban | 76.9 | 78.1 | 78.6 | 87.6 | 82.5 |
| Rural | 23.1 | 21.9 | 21.4 | 12.4 | 17.5 |
| **Per capita household income (5<sup>o</sup>)** |  |  |  |  |  |
| 1 (poorest) | 23.1 | 19.8 | 15.1 | 29.2 | 20.8 |
| 2 | 22.1 | 21.5 | 21.2 | 22.5 | 25.0 |
| 3 | 20.9 | 21.2 | 21.2 | 17.8 | 19.4 |
| 4 | 18.6 | 20.2 | 22.8 | 15.6 | 17.5 |
| 5 (richest) | 15.3 | 17.3 | 19.7 | 14.9 | 17.3 |
| **Schooling** |  |  |  |  |  |
| No schooling | 2.4 | 3.4 | 4.8 | 1.4 | 2.9 |
| Elementary school | 28.5 | 36.8 | 45.0 | 19.3 | 29.3 |
| High school | 42.6 | 35.1 | 31.4 | 40.9 | 40.3 |
| Higher education | 22.6 | 19.8 | 14.8 | 31.6 | 23.0 |
| Post graduation | 3.9 | 4.9 | 4.0 | 6.8 | 4.5 |
| **Skin color** |  |  |  |  |  |
| White/Yellow | 40.7 | 42.5 | 44.0 | 39.4 | 40.3 |
| Black/Pardo/Indigenous | 59.3 | 57.5 | 56.0 | 60.6 | 59.7 |
| **Month** |  |  |  |  |  |
| July | 20.0 | 20.3 | 21.0 | 11.9 | 25.4 |
| August | 20.3 | 19.4 | 18.8 | 16.6 | 21.0 |
| September | 20.2 | 20.1 | 20.0 | 20.7 | 18.5 |
| October | 19.7 | 20.0 | 20.1 | 23.7 | 14.9 |
| November | 19.8 | 20.2 | 20.1 | 27.1 | 20.2 |

**Table 2.**
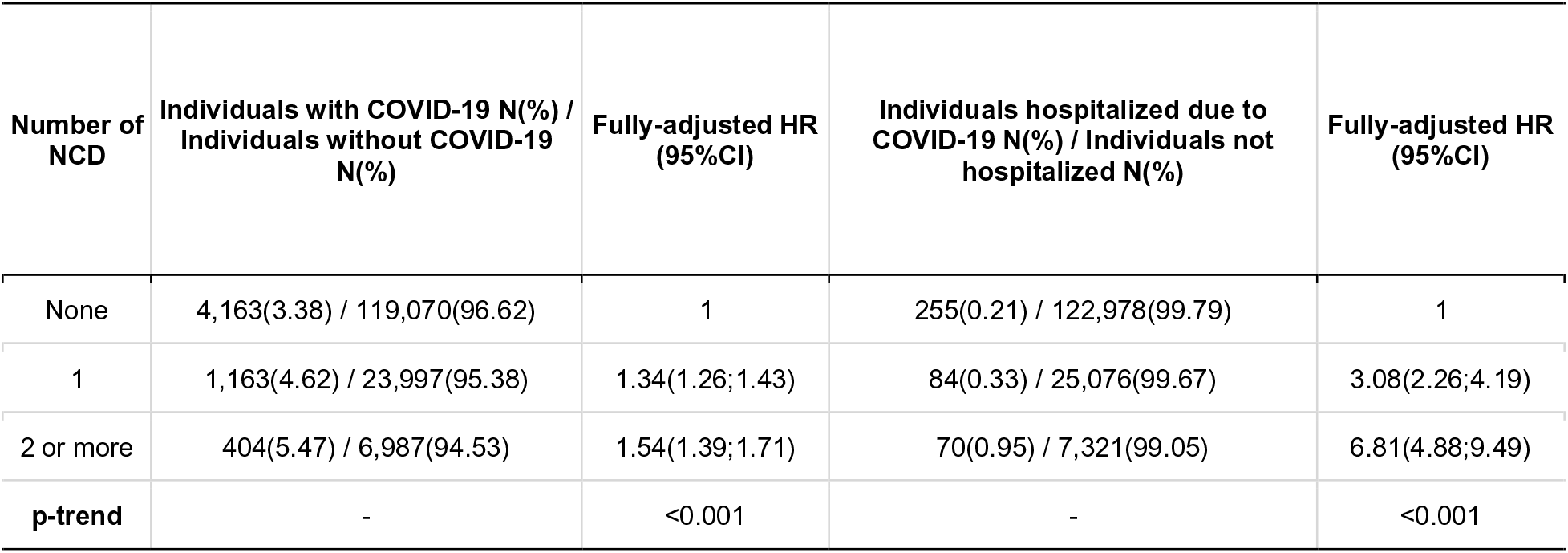
Cox models of risks of positive diagnosis and hospitalizations due to COVID-19 for Noncommunicable Diseases (NCD). Brazil PNAD COVID-19, 2020.

| Number of NCD | Individuals with COVID-19 N(%) / Individuals without COVID-19 N(%) | Fully-adjusted HR (95%CI) | Individuals hospitalized due to COVID-19 N(%) / Individuals not hospitalized N(%) | Fully-adjusted HR (95%CI) |
| --- | --- | --- | --- | --- |
| None | 4,163(3.38) / 119,070(96.62) | 1 | 255(0.21) / 122,978(99.79) | 1 |
| 1 | 1,163(4.62) / 23,997(95.38) | 1.34(1.26;1.43) | 84(0.33) / 25,076(99.67) | 3.08(2.26;4.19) |
| 2 or more | 404(5.47) / 6,987(94.53) | 1.54(1.39;1.71) | 70(0.95) / 7,321(99.05) | 6.81(4.88;9.49) |
| **p-trend** | - | <0.001 | - | <0.001 |

The stratification of the association between NCD and COVID-19 according to family income per capita is shown in Figure 1. The risk of infection by COVID-19 according to NCD tends to decrease as per capita income increases. This trend was not linear, since in the richest the risk of infection fluctuates slightly in a positive way. The risk of hospitalization for COVID-19 among NCD patients shows an inverse trend with per capita income, although this association is tenuous.

**Figure 1.**
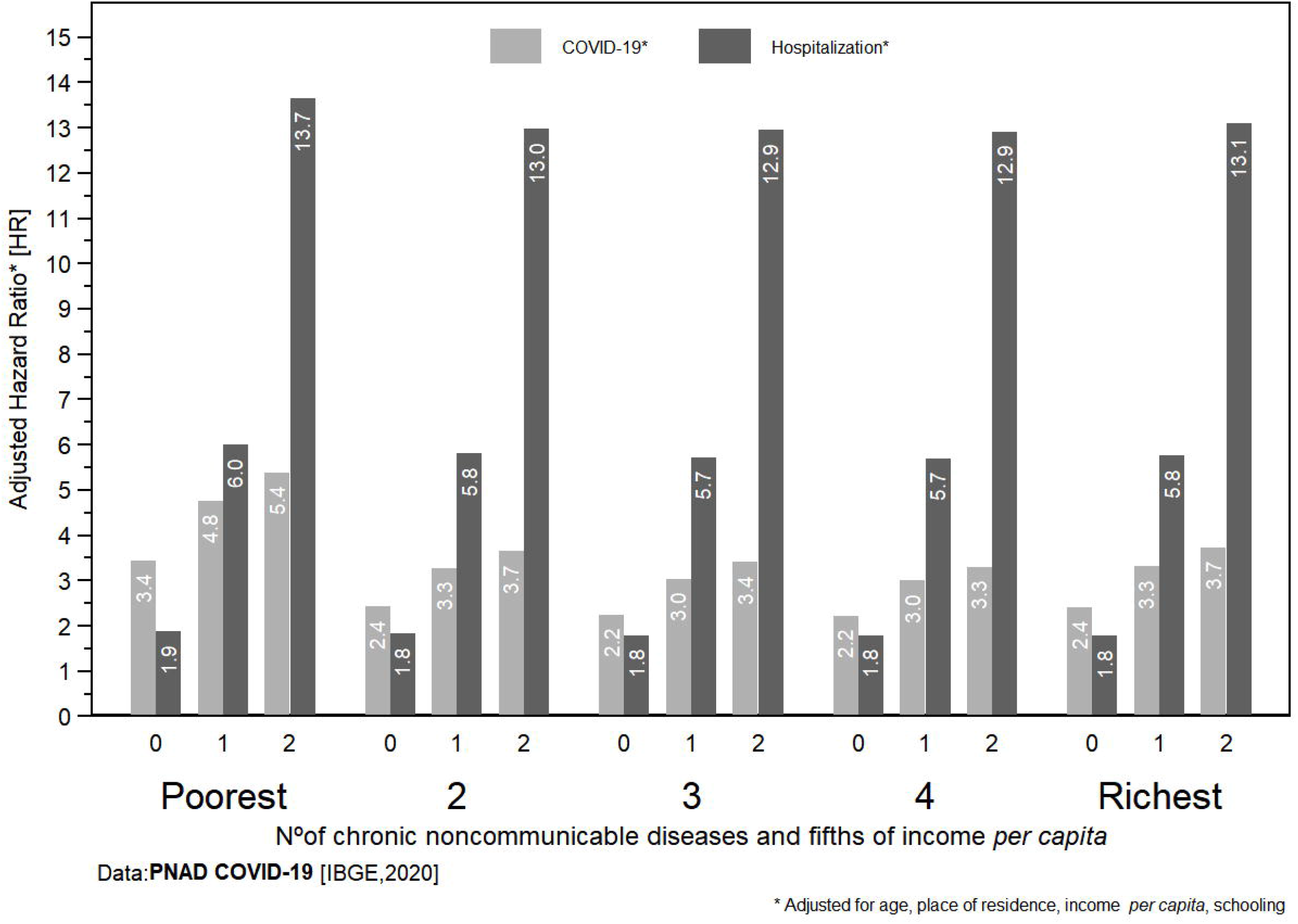
Adjusted prevalence of COVID-19 and hospitalization for COVID-19, according to the amount of NCDs and fifths of income, in Brazilian adults aged 20 to 59 years. Brazil PNAD COVID-19 (2020).

In Figure 1, we observed that the risk of hospitalization among patients without NCD did not vary according to income strata, even after adjusting for sex, age, education, income and skin color. Among patients with NCDs infected with COVID-19, the chance of hospitalization was higher among the richest than the poorest.

## DISCUSSION

Our results suggest that: a) patients with NCDs are at high risk for infection and hospitalization with the Sars-CoV-2 virus. The risk of hospitalization is approximately twice the risk of infection; b) there is a non-linear dose-response type association between the risks of infection and hospitalization due to COVID-19 and *per capita* income; c) the difference between the risks of infection or hospitalization of COVID-19 due to the burden of NCDs is non-linearly associated with the *per capita* income.

Worldwide, approximately one in three adults suffers from multiple chronic diseases (12). In Brazil, approximately 52% of the adult population has at least one chronic disease, which is responsible for 72% of the causes of death in the country (6,13).

The COVID-19 pandemic highlights the role of NCDs as risk factors for infection, hospitalization and death. Global data show that 1.7 billion people, 22% of the world population, have at least one underlying condition that puts them at increased risk for COVID-19 infection(14) In Brazil, from one third (53 million) to half (86 million) of adults have at least one risk factor for COVID-19 (7).

The association between NCDs, diagnosis and hospitalization for COVID-19, observed in our study, is in line with results from other countries (15–19). A meta-analysis carried out with Chinese data showed that the most prevalent comorbidities among patients with COVID-19 were hypertension (21%), diabetes (10%), cardiovascular disease (8%) and respiratory system disease (2%). The combined OR for presenting a severe case among those with hypertension, respiratory system disease and cardiovascular disease was 2.36 (95% CI: 1.46; 3.83), 2.46 (95% CI: 1.76; 3.44) and 3.42 (95% CI: 1.88; 6.22) respectively (15). Analyzes based on American clinical data indicate that the majority of outpatients with COVID-19 studied were African American (72%), and had at least one comorbidity (94%), including hypertension (64%), chronic kidney disease (39 %) and diabetes (38%). Severe obesity (OR = 2.0; 95% CI: 1.4; 3.6) and having chronic kidney disease (OR = 2.0; 95% CI: 1.3; 3.3) were the strongest predictors for admission to the intensive care unit (16).

Increasing evidence suggests that NCDs are determinants of the prognosis for infectious diseases (20). The gradient between the number of NCDs and the evolution to a bad prognosis of COVID-19, as found in the present study, brings up new challenges. The policy makers need to take into account the management of actions that strengthen equity attention to the social and biologically vulnerable population when elaborating strategies for health care (19).

Our results indicate that poorer individuals are more vulnerable to contagion and hospitalization due to COVID-19 in Brazil. Previous population epidemiological data (H1N1 and SARS) demonstrates the impact of social inequality as determinants for higher contagion and severity of these diseases (21–23). Factors such as lower wages, quality of public transport, unsanitary housing conditions, inefficient health care and difficulty in maintaining social isolation without significant loss of income corroborate the increased vulnerability of this population. (24,25). With regard to inequality in access to health and the needs to cope with the pandemic of COVID-19, there is an increase in the demand for health care and intensive care beds (ICU) with a range of attention to the population with dependence on public health care (3,26).

In Brazil, 71.5% of the population exclusively uses public health services, showing a direct relationship with income ranges *per capita* (27). The recommendation of WHO about the adequate ratio of Intensive Care (ICU) beds is 1 to 3 beds for every 10,000 inhabitants. In Brazil, the SUS (portuguese acronym for National Public Health System) has an average of 1.4 beds for every 10,000 inhabitants against 4.9 in private health service (28).

Our findings revealed that among NCDs patients infected with COVID-19, the chance of hospitalization was higher for the richest relative to the poorest. Income determinants seem to be associated with less access to health and intensive treatment of the disease. In our interpretation, poorer individuals, even though they are more vulnerable to the contagion of the disease, are less likely to obtain access to treatment and hospitalization for COVID-19.

As a limitation of the study, the classic limitations of observational studies can be cited, such as: self-report of information and lack of specificity and limited amount of information on clinical diagnoses. Thus, our analyzes were conditioned to the volume and type of information available in the database. As strengths of the study, longitudinal monitoring and the national representativeness of the home-based sample can be pointed out, with a high response rate. In addition, the sample size allows for consistent stratifications throughout the analysis (29).

These findings and methods of estimating the impact of vulnerable groups on the progression of the pandemic may be useful for planning and managing prevention and treatment strategies in Brazil. A better management and direction of policies that aim to obey the principle of universality of rights in health care, covering the population in a state of biological and social vulnerability is necessary. It is imperative to protect groups at greatest risk, such as people with chronic diseases and comorbidities in scenarios of broad measures of social isolation, particularly while the capacity for immunization is limited.

## CONCLUSION

The burden of NCDs increases the risk of infection and hospitalization by COVID-19. This is troublesome in a context of increasing social and health inequalities as inBrazil. This situation, associated with the prospects caused by COVID-19, projects a scenario of increased demand for clinical care in the coming years.

## Data Availability

The data used in this work can be obtained free at the Data service (https://www.ibge.gov.br/en/statistics/experimental-investigations/experimental-statistics/27975-weekly-release-pnadcovid1.html?=&t=microdados)
Further information regarding the sample design and data collection methods can be found on
the survey website (https://www.ibge.gov.br/en/statistics/experimental-investigations/experimental-statistics/27975-weekly-release-pnadcovid1.html?=&t=notas-tecnicas).

## Author Bio

Researcher and member of the Laboratory of Nutritional Assessment of Populations of the University of São Paulo. Currently study epidemiological and nutritional developments in populations.

## References

1. WHO. Weekly epidemiological update - 19 January 2021 [Internet]. 2021 Jan [cited February 18, 2021]. Available in: https://www.who.int/publications/m/item/weekly-epidemiological-update19-january-2021.

2. OMS. Oxygen sources and distribution for COVID-19 treatment centres [Internet]. 2020 [cited February 19, 2021]. Avaiable in:0020https://www.who.int/publications-detail-redirect/oxygen-sources-and-distribution-for-covid-19-treatment-centres

3. Noronha KVM de S, Guedes GR, Turra CM, Andrade MV, Botega L, Nogueira D, et al. Pandemia por COVID-19 no Brasil: análise da demanda e da oferta de leitos hospitalares e equipamentos de ventilação assistida segundo diferentes cenários. Cad Saúde Pública. 2020;36:e00115320.

4. Wu Z, McGoogan JM. Characteristics of and Important Lessons From the Coronavirus Disease 2019 (COVID-19) Outbreak in China: Summary of a Report of 72?314 Cases From the Chinese Center for Disease Control and Prevention. JAMA. 7 de abril de 2020;323(13):1239–42.

5. Malta DC, de Moura L. Probabilidade de morte prematura por doenças crônicas não transmissíveis, Brasil e regiões, projeções para 2025. 2019;13.

6. Instituto Brasileiro de Geografi a e Estatística. Pesquisa Nacional de Saúde 2019: Percepção do estado de saúde, estilos de vida, doenças crônicas e saúde bucal [Internet]. Rio de Janeiro, RJ - Brasil; 2019 [citado 18 de fevereiro de 2021]. 113 p.

7. Rezende LFM, Thome B, Schveitzer MC, Souza-Júnior PRB de, Szwarcwald CL. Adults at high-risk of severe coronavirus disease-2019 (Covid-19) in Brazil. Rev Saúde Pública. 2020;54:50–50.

8. IBGE. PNAD COVID19 – Plano amostral e ponderação. Rio de Janeiro: IBGE; 2020. 7 p.

9. Caliendo A, Hanson K. COVID-19: Diagnosis [Internet]. UpToDate. 2021 [citado 16 de março de 2021]. Disponível em: https://www.uptodate.com/contents/covid-19-diagnosis

10. Ministério da Saúde. Sobre a doença [Internet]. 2020 [citado 16 de março de 2021]. Disponível em: https://coronavirus.saude.gov.br/sobre-a-doenca

11. FDA U.S. Food & Drugs. Coronavirus Testing Basics. Coronavirus Test Basics. maio de 2020;3.

12. Roth GA, Abate D, Abate KH, Abay SM, Abbafati C, Abbasi N, et al. Global, regional, and national age-sex-specific mortality for 282 causes of death in 195 countries and territories, 1980–2017: a systematic analysis for the Global Burden of Disease Study 2017. The Lancet. 10 de novembro de 2018;392(10159):1736–88.

13. Malta DC, Moura L de, Prado RR do, Escalante JC, Schmidt MI, Duncan BB. Chronic non-communicable disease mortality in Brazil and its regions, 2000-2011. Epidemiol E Serviços Saúde. dezembro de 2014;23(4):599–608.

14. Clark A, Jit M, Warren-Gash C, Guthrie B, Wang HHX, Mercer SW, et al. Global, regional, and national estimates of the population at increased risk of severe COVID-19 due to underlying health conditions in 2020: a modelling study. Lancet Glob Health. 1o de agosto de 2020;8(8):e1003–17.

15. Yang J, Zheng Y, Gou X, Pu K, Chen Z, Guo Q, et al. Prevalence of comorbidities and its effects in patients infected with SARS-CoV-2: a systematic review and meta-analysis. Int J Infect Dis IJID Off Publ Int Soc Infect Dis. maio de 2020;94:91–5.

16. Suleyman G, Fadel RA, Malette KM, Hammond C, Abdulla H, Entz A, et al. Clinical Characteristics and Morbidity Associated With Coronavirus Disease 2019 in a Series of Patients in Metropolitan Detroit. JAMA Netw Open [Internet]. 16 de junho de 2020 [citado 22 de março de 2021];3(6). Disponível em: https://www.ncbi.nlm.nih.gov/pmc/articles/PMC7298606/

17. Xiong S, Liu L, Lin F, Shi J, Han L, Liu H, et al. Clinical characteristics of 116 hospitalized patients with COVID-19 in Wuhan, China: a single-centered, retrospective, observational study. BMC Infect Dis [Internet]. 22 de outubro de 2020 [citado 22 de março de 2021];20. Disponível em: https://www.ncbi.nlm.nih.gov/pmc/articles/PMC7578439/

18. Albashir AAD. The potential impacts of obesity on COVID-19. Clin Med. julho de 2020;20(4):e109–13.

19. Azarpazhooh MR, Morovatdar N, Avan A, Phan TG, Divani AA, Yassi N, et al. COVID-19 Pandemic and Burden of Non-Communicable Diseases: An Ecological Study on Data of 185 Countries. J Stroke Cerebrovasc Dis. setembro de 2020;29(9):105089.

20. Ogoina D, Onyemelukwe GC. The role of infections in the emergence of non-communicable diseases (NCDs): Compelling needs for novel strategies in the developing world. J Infect Public Health. 2009;2(1):14–29.

21. Tricco AC, Lillie E, Soobiah C, Perrier L, Straus SE. Impact of H1N1 on Socially Disadvantaged Populations: Systematic Review. PLoS ONE [Internet]. 25 de junho de 2012 [citado 23 de março de 2021];7(6). Disponível em: https://www.ncbi.nlm.nih.gov/pmc/articles/PMC3382581/

22. Cordoba E, Aiello AE. Social Determinants of Influenza Illness and Outbreaks in the United States. N C Med J. outubro de 2016;77(5):341–5.

23. Bucchianeri GW. Is SARS a Poor Man’s Disease? Socioeconomic Status and Risk Factors for SARS Transmission. Forum Health Econ Policy [Internet]. 22 de julho de 2010 [citado 23 de março de 2021];13(2). Disponível em: https://www.degruyter.com/document/doi/10.2202/1558-9544.1209/html

24. Carvalho L, Nassif Pires L, de Lima Xavier L. COVID-19 e Desigualdade no Brasil. 2020.

25. Michigan Department of Health and Human Services (MDHHS). COVID-19 response and mitigation strategies for racial and ethnic populations and marginalized communities [Internet]. 2020 [citado 23 de março de 2021]. Disponível em: https://www.michigan.gov/documents/mdhhs/OEMH_COVID-19_Response_Mitigation_Strategies_Targeting_Racial_Ethnic_Populations_Marginalized_Communities_FINAL_689586_7.pdf

26. Daumas RP, Silva GA, Tasca R, Leite I da C, Brasil P, Greco DB, et al. O papel da atenção primária na rede de atenção à saúde no Brasil: limites e possibilidades no enfrentamento da COVID-19. Cad Saúde Pública. 2020;36:e00104120.

27. Instituto Brasileiro de Geografi a e Estatística. Pesquisa Nacional de Saúde 2019: Informações sobre domicílios, acesso e utilização dos serviços de saúde [Internet]. Rio de Janeiro, RJ - Brasil; 2019 [cited 18 de fevereiro de 2021]. 113 p. Disponível em: https://biblioteca.ibge.gov.br/visualizacao/livros/liv101748.pdf

28. Associação de Medicina Intensiva Brasileira. Dados atualizados sobre leitos de UTI no Brasil. 2020.

29. Penna GO, Silva JAÁ, Cerbino Neto J, Temporão JG, Pinto LF. PNAD COVID-19: um novo e poderoso instrumento para vigilância em saúde no Brasil. PNAD COVID-19: a powerful new tool for public health surveillance in Brazil [Internet]. 2020 [citado 6 de abril de 2021]; Disponível em: https://www.arca.fiocruz.br/handle/icict/43323

